# Association between expedited review designations and the US or global burden of disease for drugs approved by the US Food and Drug Administration 2010–2019

**DOI:** 10.1101/2023.06.01.23290833

**Authors:** Matthew J. Jackson, Gregory Vaughan, Fred D. Ledley

**Author notes:** Corresponding author: Fred D. Ledley, Center for Integration of Science and Industry, Bentley University, Jennison Hall 144, 175 Forest Street, Waltham, MA 02452. **Mesh Headings**: “United States Food and Drug Administration” [MeSH Terms], “Global Burden of Disease” [MeSH Terms], “Drug Approval” [MeSH Terms].

## Abstract

**Introduction:** Pharmaceutical innovation can contribute to reducing the burden of disease in human populations. This research considers whether products approved by the US Food and Drug Administration (FDA) 2010–2019 and policies for expedited review of products for serious disease were aligned with the US or global burden of disease.

**Methods:** Cross-sectional study of products approved 2010–2019, their first approved indications, designations for expedited review, the burden of disease (DALYs), years of life lost (YLL), and years of life lived with disability (YLD) for 122 WHO Global Health Estimates (GHE) conditions. Statistical analyses of associations between drug approvals, disease burden of conditions comprising first approved indications, and designations for expedited review.

**Results:** The FDA approved 387 drugs 2010–2019 with lead indications for 59/122 GHE conditions. Conditions with at least one new drug had greater US DALYs (U=1193, p=0.001), US YLL (U=1144, p<0.001), global DALYs (U=1436, p=0.030), and global YLL (U=1304, p=0.004) but not US YLD (U=1583, p=0.158) or global YLD (U=1777, p=0.676). Most approvals were for conditions in the top quartiles of US DALYs or YLL, but <27% were for conditions in the top quartile of global DALYs or YLL. The likelihood of a drug having one or more expedited review designations was negatively associated (odds ratio <1) with US DALYs, US YLD, and global YLD. There was a weak negative association with global DALYs and a weak positive association (odds ratio >1) with US and global YLL.

**Conclusions:** Drug approvals 2010–2019 were more strongly aligned with US than global disease burden and more strongly associated with YLL than YLD. Expedited review pathways were not aligned with the US or global burdens of disease and prioritize YLL over YLD. These results may inform policies to incentivize pharmaceutical innovation better aligned with global burden of disease.

**KEY QUESTIONS:** 

**What is already known on this topic:** Pharmaceutical innovation is strongly influenced by (US) market opportunities and poorly aligned with the global burden of disease. Previous studies have suggested that regulatory policies designed to expedite development of products for serious disease could promote better alignment between pharmaceutical innovation and global disease burdens.

**What this study adds:** Drug approvals by the US Food and Drug Administration 2010–2019 were more strongly associated with the US than global burden of disease and were disproportionately focused on disorders contributing to premature death as opposed to disability. The odds of a product being designated for expedited review was negatively associated with the burden of disease and measures of disability but positively associated with years of life lost to disease.

**How this study might affect research, practice, or policy:** This work demonstrates a persistent failure of drug development for conditions that contribute the most to the global burden of disease and disabilities that is not addressed with policies for expedited review. This analysis may inform new policy explicitly designed to promote innovation for indications associated with the greatest disease burden and, specifically, the burden associated with disabilities.

## INTRODUCTION

This study explores the relationships between the burden of disease in US and global populations, the drugs approved by the US Food and Drug Administration (FDA) 2010–2019, and the US expedited review pathways designed to promote development of drugs for “serious diseases.” Previous studies have suggested that the number of drug approvals in different therapeutic areas is generally aligned with their burden of disease in the US^1^ but not their contribution to global disease burden.^1–6^

Pharmaceutical R&D spending and development is typically associated with the size of the available market for different indications and the anticipated returns on investment.^7–12^ In this context, it has been argued that industry’s underinvestment in research and development related to diseases that are more prevalent outside of the US^5 13^ and the corresponding paucity of products addressing the morbid diseases of global populations represents a classic market failure in which the pharmaceutical industry considers the market for such products inadequate to justify the cost of investment.^14–16^ While alternative business models involving philanthropic and non-governmental organizations have had notable success,^16–18^ it is classically the role of government to rectify such market failures through regulatory or economic policy.^19^

Over the past 40 years, a series of regulations in the US and EU have been implemented to redress analogous market failures involving rare (orphan) diseases, paediatric disorders, and serious diseases with characteristics that make investment in these products unattractive.^20–23^ The prototype for such regulations was the US Orphan Drug Act of 1983,^24^ which was dramatically successful in promoting development of drugs for rare diseases,^25 26^ though the Act also had unintended consequences related to drug pricing, the adequacy of clinical trials in small populations, and widespread applications to precision medicine.^27^ Subsequently, four expedited review programs were legislated (“fast track”, “breakthrough therapy”, “accelerated approval”, and “priority review”) to incentivize development of products for serious diseases and address significant unmet medical needs, provide substantial improvement over existing therapies, require use of surrogate endpoints to demonstrate clinical efficacy, offer significant improvement in safety or effectiveness, include paediatric studies, or address specific infectious diseases.^28–30^ These pathways offer industry the opportunity for greater returns on investment by reducing the net cost or timeline of product development or increasing returns through government-granted monopolies. While questions have been raised about the impact on health outcomes, safety, and innovativeness of products receiving expedited review,^28 31–33^ these programs have been widely utilized to the point that the majority of new drugs coming to market have at least one expedited review designation.^22^

It has been proposed that such regulations could provide a path for promoting development of drugs for the morbidities of global populations.^34–36^ In fact, elements of the priority review program are explicitly aimed at promoting drug development for a list of tropical diseases.^37 38^ While a number of drugs for tropical disease have been approved through priority review,^39 40^ there is insufficient evidence to assess the potential impact of this approach.^37 41^

### Objectives

The objective of this study was to ask whether the drugs approved by the US FDA through the decade 2010–2019 (prior to COVID) were aligned with measures of the US or global burden of disease at the outset of the decade in 2010 and whether this alignment was promoted by the four expedited review pathways in the US. Specifically, this analysis identified the drugs approved by the US FDA, the first approved indications for these products, the Global Health Estimates (GHE) conditions associated with these indications, metrics of the US and global burden of disease associated with these conditions, and designations for expedited review. The analysis describes the relationship between the number of product approvals for indications in each condition and its contribution to the US or global disease burden. The analysis further estimates the relative likelihood (odds ratio) of a product being granted one or more designations for expedited review based on metrics of the disease burden for the condition comprising the first approved indication.

The burden of disease in whole populations is classically measured in Disability-Adjusted Life Years (DALYs).^42 43^ A DALY is calculated as the sum of Years of potential life lost (YLL) and Years of healthy life lost to disability (YLD). YLL measures the proportion of the burden of disease associated with premature mortality calculated from the age of death to relative life expectancy. YLD measures the proportion of the burden of disease associated with morbidity, calculated as the product of the Disability Weight of a condition and its prevalence in a population.^44 45^ This analysis considered each of these measures of disease burden independently.

The results confirm previous observations that the number of drug approvals is more closely associated with the US rather than the global burden of disease measured by DALYs.^1–6^ Analyses from the present study further show that this association is largely limited to measures of premature mortality (YLL) rather than morbidity (YLD). The analysis also shows that the likelihood of a drug having one or more designations for expedited review is negatively associated with the burden of disease (DALYs), with a positive association between expedited approvals and measures of mortality (YLL) negated by a strong negative association with measures of morbidity (YLD). These results are discussed in the context of the continuing challenge of achieving global equity in pharmaceutical innovations that address humankind’s most burdensome diseases.

## METHODS

### Study design

This cross-sectional study describes associations between the lead indications for drugs approved by the FDA 2010–2019, designations of these products for expedited review, and WHO GHE metrics for the US and global burden of disease of conditions comprising these indications.

### Data sources

FDA-approved products 2010–2019 (NDA or BLA) and dates of first approval were identified from annual FDA reports (CDER: https://www.fda.gov/drugs/development-approval-process-drugs/new-drugs-fda-cders-new-molecular-entities-and-new-therapeutic-biological-products; CBER: https://www.fda.gov/vaccines-blood-biologics/development-approval-process-cber/2022-biological-approvals). Products derived from blood or tissue, diagnostic agents, vaccines, and antimicrobials were excluded. Expedited and orphan designations for products approved by CDER were identified in the CDER annual reports, or by accessing the individual Summary Basis for Regulatory Action on that product.

The first approved indication was identified in the “Full Prescribing Information” (product label) for each product (CDER: https://www.accessdata.fda.gov/scripts/cder/daf/index.cfm, CBER: https://www.fda.gov/vaccines-blood-biologics/development-approval-process-cber). The first approved indication was matched to corresponding ICD-10 codes via the WHO ICD-10 lookup tool (https://icd.who.int/browse10/2016/en). Drugs were attributed to a GHE condition by matching the ICD-10 code to the corresponding GHE ICD-10 code cluster (https://ghdx.healthdata.org/record/ihme-data/gbd-2019-cause-icd-code-mappings).^46^ For certain analyses, GHE conditions were categorized into 13 therapeutic areas adapted from the WHO Health Observatory data repository disease hierarchy.^47^

DALYs, YLL, and YLD were obtained for all 132 available GHE conditions for the year 2010 from the WHO Health Observatory data repository GHE-2020.^48^ Burden estimates for 10 conditions in the categories of “Sudden infant death syndrome” and “Injuries” were excluded.

The final burden dataset consisted of 122 GHE conditions, 59 of which had at least one associated FDA-approved drug 2010–2019.

### Statistical methods

Differences between the burden of disease metrics for conditions with one or more drug approvals or no drug approvals were analysed with Mann-Whitney tests. Significance is inferred with p<0.05. Bonferroni corrections of 8 were applied to account for multiple testing, with a corrected p<0.0065 corresponding to the p=0.05 threshold.

Probability estimates of a product receiving expedited designations (dependent variable) as a function of the DALYs, YLL, or YLD for the GHE condition associated with its indication (independent variable) were assessed using univariate binary logistic regression models (Model 1):

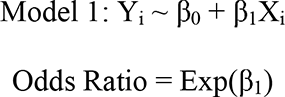

where Y_i_ indicates designation or no designation for a drug *i*, β_0_ represents the Y intercept, β_1_ represents the slope coefficient, X_i_ represents the independent variable (DALYs, YLL, or YLD) of the disease corresponding to drug *i*, and Exp(*x*) is *e^x^*. Analysis was performed independently for each expedited designation and metric of disease burden. Model 1 was used for datasets comprising all drugs, drugs indicated for neoplastic disease, and drugs indicated for non-neoplastic disease.

Mann-Whitney tests were performed comparing ranked DALYs, YLL, or YLD for neoplasm versus non-neoplasm GHE conditions. Mann-Whitney and logistic regression were performed in SPSS Statistics version 27 (IBM). All other data manipulation and analysis was performed in Excel.

This study did not involve research on human subjects and was exempt from ethics review under the Declaration of Helsinki. This manuscript follows STROBE guidelines where applicable.

## RESULTS

### Drug approvals, expedited review designations, and first approved indications

The FDA approved 387 drugs 2010–2019 excluding vaccines and biological products derived from blood or tissues (supplemental table 1). Of these, 227/387 (58.7%) were granted at least one designation for expedited review including 49 (12.7%) accelerated approvals, 78 (20.2%) breakthrough therapies, 142 (36.7%) fast tracks, and 207 (53.5%) priority reviews. Across all approved products, there was an average of 1.2 designations for expedited review. These fractions are consistent with observations by others (table 1).^31 49 50^

**Table 1.** Number of drugs approved by the FDA 2010–2019 and designations for expedited review by therapeutic area.

|  | Drugs | Accelerated | Breakthrough | Fast track | Priority | Average | >1 |
| --- | --- | --- | --- | --- | --- | --- | --- |
| Therapeutic area | # | # drugs (% drugs in class) |  |  |  |  |  |
| Neoplasms | 97 | 38 (39.2%) | 39 (40.2%) | 46 (47.4%) | 80 (82.5%) | 2.1 | 90 (92.8%) |
| Endocrine, blood, immune disorders | 73 | 3 (4.1%) | 16 (21.9%) | 28 (38.4%) | 37 (50.7%) | 1.2 | 39 (53.4%) |
| Neurological & sense organ | 50 | 2 (4%) | 7 (14%) | 11 (22%) | 22 (44%) | 0.8 | 22 (44%) |
| Infectious and parasitic diseases | 37 | 2 (5.4%) | 10 (27%) | 22 (59.5%) | 27 (73%) | 1.7 | 28 (75.7%) |
| Digestive & genitourinary | 31 | 1 (3.2%) | - | 13 (41.9%) | 16 (51.6%) | 1.0 | 16 (51.6%) |
| Respiratory | 18 | - | 2 (11.1%) | 5 (27.8%) | 5 (27.8%) | 0.7 | 6 (33.3%) |
| Skin & oral | 16 | - | 1 (6.3%) | 2 (12.5%) | 4 (25%) | 0.4 | 4 (25%) |
| Cardiovascular | 16 | 1 (6.3%) | - | 5 (31.3%) | 7 (43.8%) | 0.8 | 8 (50%) |
| Musculoskeletal | 13 | - | - | 2 (15.4%) | 2 (15.4%) | 0.3 | 2 (15.4%) |
| Diabetes mellitus | 12 | - | - | - | 1 (8.3%) | 0.1 | 1 (8.3%) |
| Mental and substance use | 12 | - | 1 (8.3%) | 2 (16.7%) | 2 (16.7%) | 0.4 | 3 (25%) |
| Maternal, neonatal, & congenital | 4 | - | - | 1 (25%) | - | 0.3 | 1 (25%) |
| Other | 8 | 2 (25%) | 2 (25%) | 5 (62.5%) | 4 (50%) | 1.8 | 7 (87.5%) |
| **Total** | **387** | **49 (12.7%)** | **78 (20.2%)** | **142 (36.7%)** | **207 (53.5%)** | **1.2** | **227 (58.7%)** |

FDA approved drugs and expedited designations for each drug are shown in supplemental table 1.

The first approved indications for products in this dataset were associated with 188 unique ICD-10 codes (supplemental table 1) corresponding to 59/122 (48%) GHE conditions (supplemental table 2). The GHE conditions were further categorized into 13 therapeutic areas (table 2). The largest fraction of drug approvals was for “neoplasms” (n=97/387) followed by “Endocrine, blood & immune disorders” (n=73/387), and “neurological & sense organ” (n=50/387). Drugs for infectious and parasitic disease included treatments for hepatitis C, HIV, malaria, Chagas disease, leishmaniasis, fascioliasis, onchocerciasis, and tuberculosis, diseases of particular concern in the tropical and developing world. Table 2 shows the US and global burden of disease for all 122 GHE conditions categorized by therapeutic area measured by DALYs, YLL, and YLD. Notably, the US burden estimates represent <6% of global DALYs, YLL, and YLD.

**Table 2.**
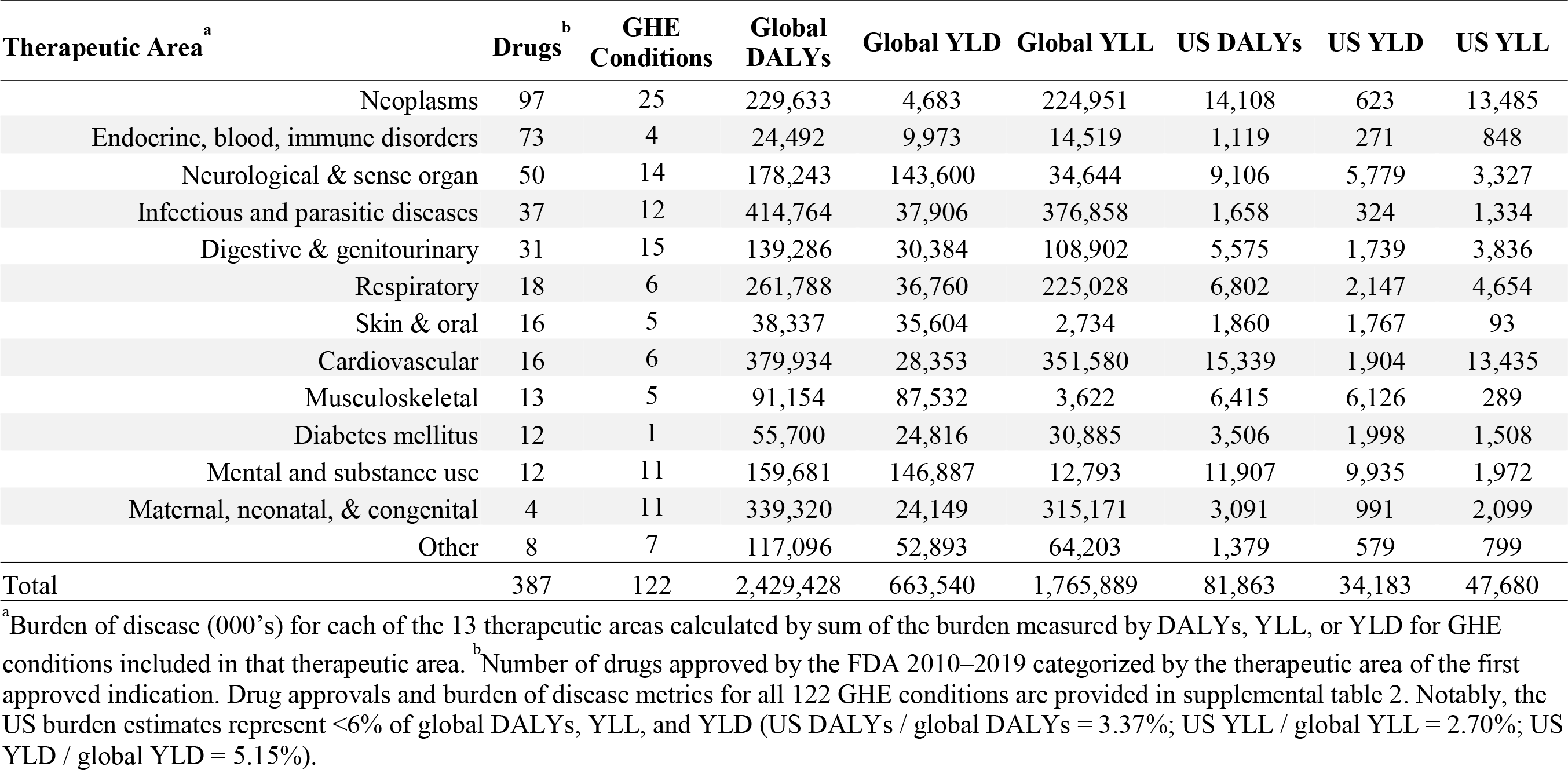
US and global burden of disease estimates (‘000s) by therapeutic area as measured by DALYs, YLL, and YLD (GHE-2020)

Products indicated for neoplasm were most likely to have expedited designations, with 90/97 (92.8%) having at least one expedited designation and an average of 2.1 designations per product. Products for infectious and parasitic diseases were also likely to have expedited designations, with 28/37 (75.7%) having at least one expedited designation and an average of 1.7 designations per product (table 1). Seven drugs were indicated for diseases explicitly listed by the FDA as tropical diseases eligible for priority review (supplemental table 3). Therapeutic areas with the lowest average number of expedited designations were diabetes mellitus (0.1 designations per product), maternal, neonatal, and congenital (0.3 designations per product), musculoskeletal (0.3 designations per product), and mental and substance use (0.4 designations per product) (table 1).

### Associations between drug approvals and US or global burden of disease

Table 3 compares US and global DALYs, YLL, and YLD for 59 conditions associated with at least one new drug approval versus 63 conditions with no approval. US DALYs and YLL were significantly higher for conditions with at least one new drug approval (US DALYs, U=1193, p=0.001; US YLL, U=1144, p<0.001). Global DALYs and YLL were also higher for conditions with at least one new drug approval (global DALYs, U=1436, p=0.030; global YLL, U=1304, p=0.004), though the difference in global DALYs was not significant after Bonferroni correction.

**Table 3.** Mann-Whitney analysis of differences in disease burden between GHE disease states with or without drug <u>approvals 2010–2019</u>

| Burden metric | Approved Drug | All GHE disease states |  |  |  |  | Non-neoplasm GHE disease states |  |  |  |  |
| --- | --- | --- | --- | --- | --- | --- | --- | --- | --- | --- | --- |
|  |  | N | Median ('000s) | Mean Rank | Mann-Whitney U | Asymp. Sig. (2-tailed) | N | Median ('000s) | Mean Rank | Mann-Whitney U | Asymp. Sig. (2-tailed) |
| Global DALYs | No | 63 | 7212 | 54.8 | 1436 | 0.030 | 53 | 7255 | 43.7 | 883 | 0.040 |
|  | Yes | 59 | 11197 | 68.7 |  |  | 44 | 13503 | 55.4 |  |  |
| Global YLD | No | 63 | 1741 | 60.2 | 1777 | 0.676 | 53 | 3127 | 46.7 | 1044 | 0.377 |
|  | Yes | 59 | 1341 | 62.9 |  |  | 44 | 3950 | 51.8 |  |  |
| Global YLL | No | 63 | 1129 | 52.7 | 1304 | 0.004 | 53 | 812 | 43.0 | 846 | 0.020 |
|  | Yes | 59 | 6203 | 70.9 |  |  | 44 | 6196 | 56.3 |  |  |
| US DALYs | No | 63 | 143 | 50.9 | 1193 | 0.001 | 53 | 132 | 41.4 | 761 | 0.003 |
|  | Yes | 59 | 523 | 72.8 |  |  | 44 | 513 | 58.2 |  |  |
| US YLD | No | 63 | 47 | 57.1 | 1583 | 0.158 | 53 | 57 | 45.0 | 955 | 0.126 |
|  | Yes | 59 | 70 | 66.2 |  |  | 44 | 124 | 53.8 |  |  |
| US YLL | No | 63 | 15 | 50.2 | 1144 | <0.001 | 53 | 8 | 40.7 | 724 | 0.001 |
|  | Yes | 59 | 287 | 73.6 |  |  | 44 | 114 | 59.0 |  |  |

Significance is inferred with a Bonferroni correction of 8, such that a calculated p value of 0.0065 is equivalent to a threshold of 0.05.

There was no significant difference in the YLD for conditions with drug approvals and without an approved drug (US YLD, U=1583, p=0.158; global YLD, U=1777, p=0.676).

Furthermore, when the analysis included all 122 conditions with or without approvals was repeated using non-neoplastic conditions, US DALYs and YLL remained significantly higher for conditions with at least one approval (US DALYs, U=761, p=0.003; US YLL, U=724, p=0.001). No significant difference was seen between conditions with or without at least one drug approval across any of the global burden of disease measures (table 3).

Figure 1 illustrates the relationship between US and global DALYs for the 122 GHE conditions ranked by US DALYs, the boundaries of quartiles 1 to 4 of US to global DALYs, and the number of new drugs for indications in each condition. While the US and global DALYs are highly correlated (R=0.615, p<0.001), there are also substantive differences (supplemental table 4). For example, multiple neoplasms, cardiovascular diseases, drug and alcohol use disorders, skin diseases, digestive diseases, osteoarthritis, and neurological conditions were in Q4 (the top quartile) of the US disease burden but were in Q2 or Q3 of the global burden. Conversely, tuberculosis, meningitis, maternal conditions, rheumatic heart disease, and parasitic and vector diseases were in Q1 (the lowest quartile) of the US disease burden but were in Q3 or Q4 of the global burden (supplemental table 2). Figure 1 also illustrates the number of drug approvals for indications in each condition, illustrating the greater number of drugs for conditions in the highest quartiles of US disease burden and the distribution of these conditions across all four quartiles of the global burden.

**Fig 1.**
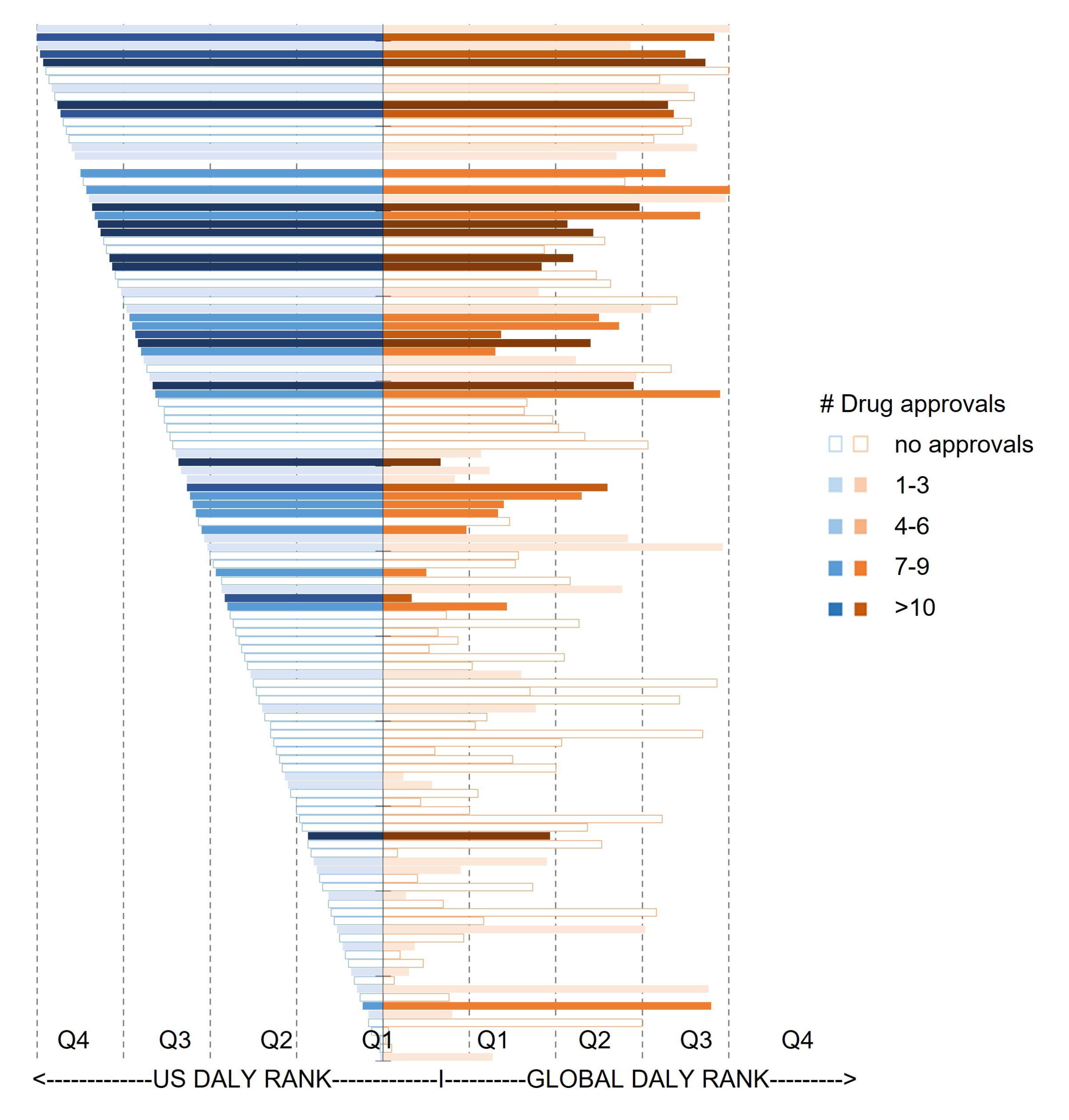
Relationship between FDA drug approvals and the US or global burden of disease of GHE conditions associated with their lead indications. The 122 GHE conditions in the GHE- 2020 study were ranked by US DALYs. Bars are scaled by US DALYs (left, blue) or global DALYs (right, orange). Bars are colored by the number of products approved by the FDA 2010– 2019 with first approved indications for that condition. Empty bars indicate no approvals. Darker bars indicate larger numbers of approvals. Gridlines indicate quartiles of DALYs with Q4 representing disorders with the highest burden and Q1 representing the lowest burden.

Figure 2A shows the number of drugs for conditions in each quartile of disease burden measured by DALYs, YLL, or YLD for both US and global populations (data breakdown in supplemental table 5). The majority of drug approvals were for diseases indicated for conditions in the top quartile of US DALYs. This was not evident for the global burden of disease, where <27% of products were approved for conditions in the top quartile (Q4) of global DALYs, YLL, or YLD (figure 2A, supplemental table 5). Both are consistent with the Mann-Whitney analysis observations that conditions with at least one drug approval had significantly greater ranked US DALYs and YLL than conditions without approved products, while conditions with at least one drug approval had lower ranked global burden of disease metrics than those without an approved product (table 3).

**Fig 2.**
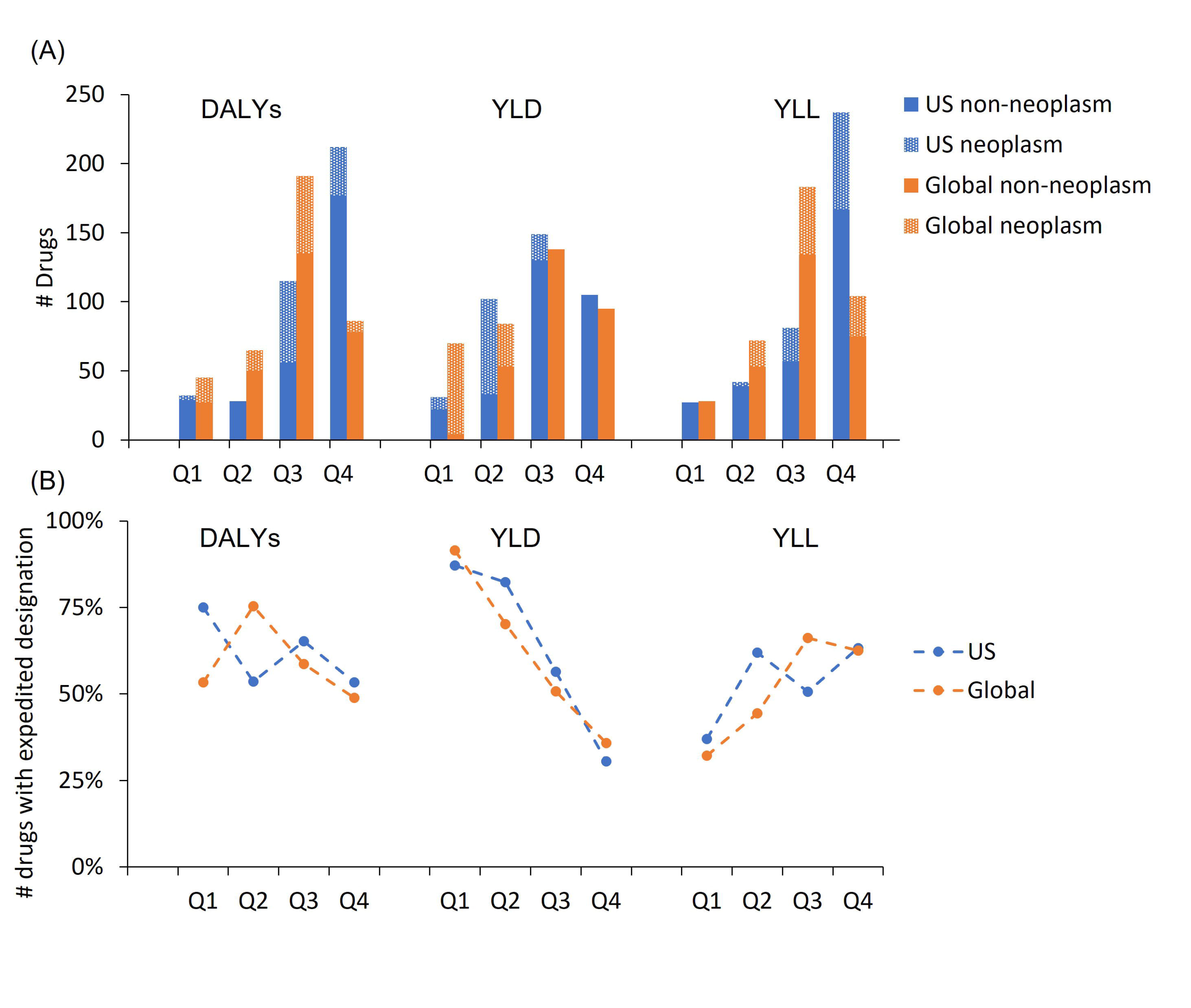
Association of FDA drug approvals and designations for expedited review across quartiles of US and global burden of disease. (A) Number of FDA drug approvals 2010–2019 is shown quartiles of the US (blue) or global (orange) burden of disease for GHE conditions comprising the lead indication. The burden of disease is measured by DALYs, YLL, or YLD. The number of FDA approvals for non-neoplastic conditions is shown in patterned fill. (B) The proportion of drugs approved 2010–2019 with at least one expedited designation across quartiles of disease burden. The burden of disease for 122 GHE conditions is measured by DALYs, YLL, and YLD and was divided by quartile with Q1 representing the lowest burden and Q4 representing the highest burden.

These analyses demonstrate a strong association between drug approvals and the US burden of disease measured by DALYs or YLL and a demonstrably weaker association between drug approvals and global disease metrics. These analyses also show that these associations are largely limited to measures of premature mortality (YLL) and that there is little association between drug approvals and measures of disability (YLD).

Figure 2A also illustrates differences in the association of drugs for neoplastic disease and non-neoplastic disease and quartiles of US or global disease burden. Drugs for neoplasms are predominantly associated with conditions in the top two quartiles of US DALYs and YLL, the lowest two quartiles of US and global YLD, and widely across quartiles of global DALYs and YLL. These results suggest that the large number of approvals for neoplastic indications contributes to the overall absence of an association between drug approvals and either US or global YLD. This is particularly pronounced for global YLD, where most neoplastic conditions are in the lowest quartile (figure 2A). Nevertheless, there were fewer products for conditions in the top quartile of both US and global YLD than for conditions in the third quartile.

### Association between expedited review designations and burden of disease

Figure 2B shows the relationship between the fraction of approvals having one or more expedited designations and the quartiles of US or global DALYs, YLL, or YLD. Figure 3A,C shows the calculated odds ratios of a product having at least one designation for expedited review as a function of US or global burden of disease metrics associated with the condition comprising the lead indication.

**Fig 3.**
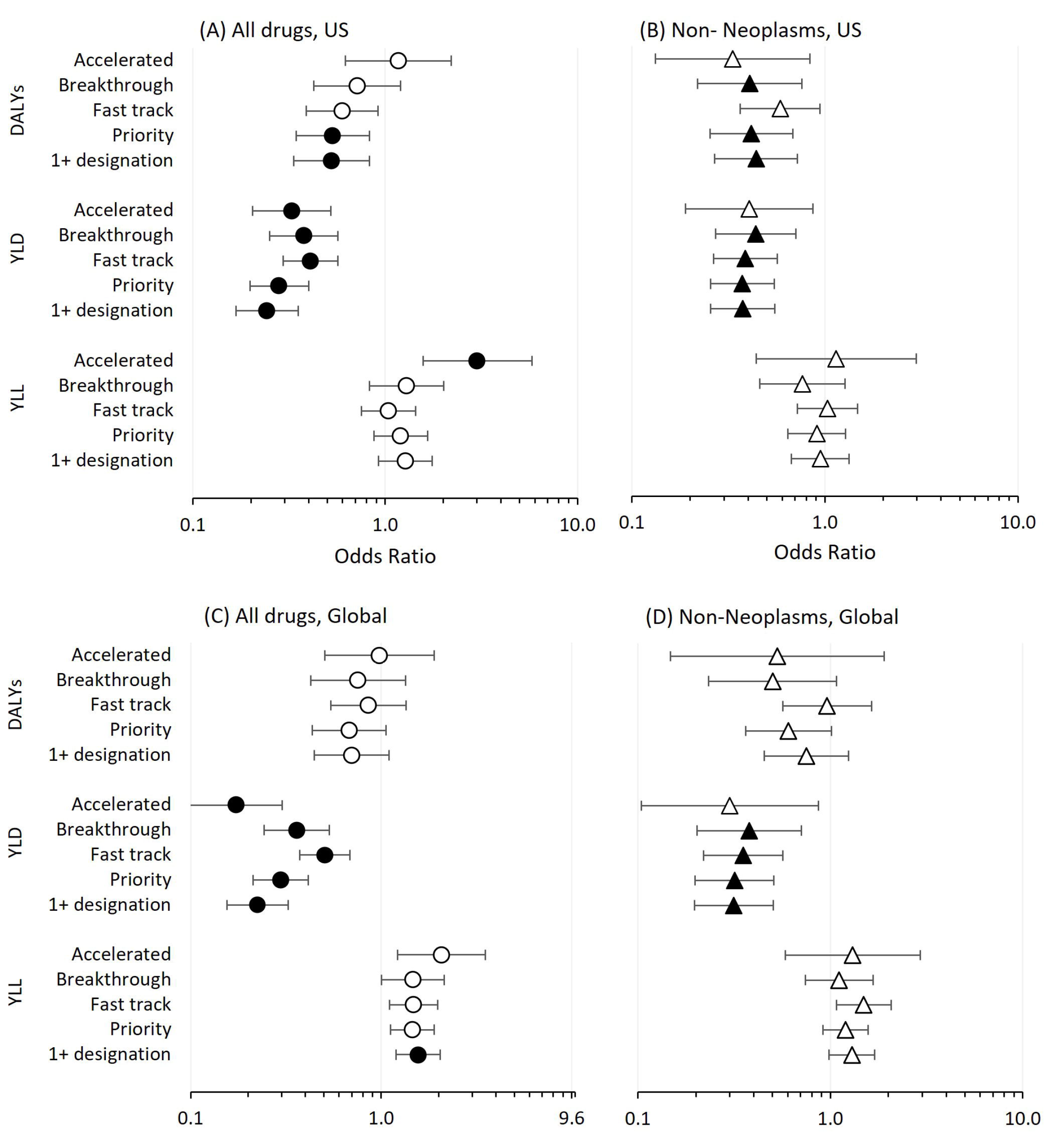
Odds ratios representing the likelihood of receiving an expedited designation as a function of burden of disease. The odds ratio of approved drugs having a designation for expedited review was calculated as a function of burden of disease metrics for GHE conditions comprising the first approved indication. Data are shown for US or global burden of disease metrics including DALYs, YLL, or YLD. (A) Odds ratio calculated for 387 products approved 2010–2019 and US burden of disease metrics. (B) Odds ratio calculated for 290 products indicated for non-neoplastic conditions and US burden of disease metrics. (C) Odds ratio calculated for 387 products approved 2010–2019 and global burden of disease metrics. (D) Odds ratio calculated for 290 products indicated for non-neoplastic conditions and global burden of disease metrics approved 2010–2019. Data are shown as the average and 95% confidence interval. Filled symbols represent data with p<0.0065 corresponding to the significance threshold of p=0.05 after a Bonferroni correction of 8 to account for multiple testing.

There was a negative trend in the fraction of drugs with at least one expedited review designation across quartiles for either US or global DALYs (figure 2B). This trend was also evident with odds ratios significantly <1 for a product having at least one expedited review designation as a function of US DALYs (OR=0.526, 95% C.I. 0.335 to 0.826, p=0.005) (figure 3A) and a negative association with global DALYs (OR=0.673, 95% C.I. 0.428 to 1.057, p=0.085) (figure 3C).

There was a positive trend in the fraction of drugs having at least one expedited designation across quartiles of either US or global YLL (figure 2B). This was also evident by the odds ratio of >1 for a product having at least one designation for expedited review as a function of US or global YLL (US: OR=1.274, 95% C.I. 0.924 to 1.758, p=0.140; global: OR=1.501, 95% C.I. 1.150 to 1.958, p=0.003) (figures 3A,C).

In contrast, there was a negative trend in the fraction of drugs having at least one expedited designation across quartiles of US or global YLD (figure 2B). This trend was also evident in the statistically significant odds ratios <1 for a product having at least one expedited designation as a function of US or global YLD (US: OR=0.243 95% C.I. 0.168 to 0.352, p<0.001; global: OR=0.215, C.I. 0.149 to 0.311, p<0.001) (figure 3A,C). Odds ratios for US and global expedited designations are shown in figure 3 and full binary logistic regression variables are shown in supplemental table 6. Odds ratios for each individual designation were similar to odds ratios for having at least one expedited designation (figure 3A,B,C,D).

Similar patterns were observed when considering the 290 drugs approved across 97 non-neoplastic conditions (figure 3B,D). For these drugs, the odds ratio of having at least one expedited review designation as a function of US DALYs was significantly <1 and not significantly <1 as a function of global DALYs (US: OR=0.440 95% C.I. 0.269 to 0.719, p=0.001; global: OR=0.751, C.I. 0.454 to 1.243, p=0.265). As a function of US and global YLD, the odds ratio of non-neoplastic drugs having at least one expedited review designation was significantly <1 (US: OR=0.375 95% C.I. 0.098 to 0.131, p<0.001; global: OR=0.315, C.I. 0.197 to 0.505, p<0.001).

Analysis of the 97 products indicated for neoplastic disease was limited by the paucity of products without expedited designations and, consequently, large standard errors. These results are shown in supplemental table 6.

## DISCUSSION

This research examined the association between the first approved indication for drugs approved by the FDA 2010–2019 and the US or global burden of disease for the associated GHE disease condition. Specifically, this analysis asked two questions; first, whether drug approvals for each GHE condition were related to the US or global burden of disease measured by DALYs, YLL or YLD; and second, whether products with indications having greater US or global burden of disease were more likely to be designated for expedited review and eligible for the incentives designed to promote development of products for serious diseases.

### Drug approvals and the US or global burden of disease

These analyses demonstrated a strong association between drug approvals and conditions with higher US DALYs. This was evident both in the observation that conditions with at least one new drug approval had significantly higher US DALYs than those without new approvals, and that the majority of all drug approvals had an indication associated with conditions in the top quartile of US DALYs. There was a less pronounced difference in the global burden of disease for conditions with or without new drug approvals, with <23% of drug approvals indicated for conditions ranking in the top quartile of global disease burden. These results are consistent with previous studies showing that drug approvals are more closely associated with the US burden of disease than the global burden of disease.^1 2 4–6 51^ It is noteworthy that the global burden of disease represents >96% of the total DALYs associated with the 122 GHE conditions. Thus, these results suggest there is little relationship between drug approvals and the composite health needs of global human populations.

An unexpected finding in this work was the disparity between the strong association observed between the number of drug approvals and the disease burden associated with premature death (YLL) and the absence of any association between the number of drug approvals and years of life lived with disability (YLD). While this disparity is partially related to the large number of drugs for neoplastic conditions with high YLL and proportionally lower YLD, the disparity was still observed when considering only drugs for non-neoplastic conditions.

The underrepresentation of drugs indicated for conditions with the highest burden of disability is paradoxical given that the burden of disability measured by US YLD represents 42% of the total US disease burden measured by US DALYs. Evidence suggests that new drug launches can significantly reduce broad measures of disability^52^ and that individuals with disabilities filled an average of five times more prescriptions than those without disabilities,^53^ generating sales representing 32% of the US pharmaceutical market^53^ even with the relatively low fraction of new products indicated for conditions with greater years of life lived with disability.

### Expedited review designations and the burden of disease

It has been proposed that pathways for expedited review of products for serious disease could incentivize alignment with US or global burdens of disease.^34 35^ The present analysis does not support this association. On the contrary, this work shows that the likelihood of a drug having at least one designation for expedited review, priority review, or fast track review was generally lower for drugs with indications for conditions having a greater US or global burden of disease as measured by either US or global DALYs. The negative association between designations for priority review and measures of global disease burden is particularly notable in that this designation specifically includes a list of tropical diseases of particular concern to global populations (figure 3C).^37 38^ This analysis also showed distinctly different associations of YLL and YLD with the likelihood of having a designation for expedited review. While there was a weak, positive association between the likelihood of a product having a designation for expedited review and the burden of premature death measured by YLL, there was strong, negative association with the burden of chronic disease measured by YLD. This dynamic might contribute to the observed paucity of products indicated for conditions with the highest level of disability relative to those indicated for conditions with greater years of life lost. By disproportionately reducing the timelines or costs of developing products for conditions with high YLL relative to those with greater YLD, these policies may differentially increase the potential returns on investment in products addressing premature death and make investments in products for treating long-term morbidities less attractive.

## Limitations

First, the study design focuses explicitly on the first approved indication for drugs in the dataset, many of which receive supplemental indications over time. Thus, the study may underestimate the impact of products that are ultimately approved for multiple applications or have substantial off-label use. Second, the study considers the burden of disease at the level of GHE conditions, each of which comprises multiple indications based on their ICD-10 codes. The study may, thus, overestimate the burden of disease associated with any one indication. Third, this work looks explicitly at the burden of disease measured by WHO GBD metrics and may not fully account for the economic or social costs of disability or disease or the burdens associated with co- morbidities arising from a disorder or its treatment. Fourth, this analysis only considered drugs approved by the US FDA. There is no evidence that these findings apply to drugs approved only in nations outside the US.

## Conclusion and policy implications

This work demonstrates a persistent failure of pharmaceutical innovation for conditions contributing to the greatest global burden of disease. This failure has been attributed to the pharmaceutical industry’s primary focus on developing products for large markets,^7–12^ primarily those in the US that offer the greatest return on investment. It has been suggested that the existing regulations for fast track, breakthrough therapy, accelerated approval, or priority review, which were designed to address some of the resulting market failures, might also incentivize development of products for conditions with the greatest burden of disease. The present analysis suggests that this is not being achieved.

Instead, this analysis shows that conditions with the highest global burden of disease are less likely to have designations for expedited review. These data also suggest that expedited review may differentially incentivize products for conditions associated with greater years of life lost (YLL) over those associated with greater disability (YLD). Incentivizing industry to address the need for drugs that address the years of life lived with disability may require new regulatory mechanisms beyond those focused on near death conditions. Further research needs to be directed at refining metrics for the burden of disease to describe individual conditions as well as modelling the impact of potential regulatory regimens on pharmaceutical innovation.

## Data Availability

All databases used in writing this article are publicly available and cited in the text. All extracted data is provided in supplemental table 1 or supplemental table 2. Results from statistical outputs are provided in supplemental table 4-6.

## ACKNOWLEDGEMENTS

We would like to thank Dr. Michael Boss, Dr. Nancy Hsiung, and Bruce Leicher, Esq., for advice and critical reading of the manuscript as well as Juliana Harrison, M.B.A., for managing and editing this submission.

## Article Information

### Corresponding Author

Fred D. Ledley, M.D., Center for Integration of Science and Industry, Bentley University, Jennison Hall 144, 175 Forest Street, Waltham, MA 02452

### Contributors

FDL contributed as corresponding author. All authors designed the study. All authors analysed the data and performed the statistical analyses. MJJ and FDL drafted the initial manuscript. FDL is the guarantor. All authors reviewed the drafted manuscript for critical content. All authors approved the final version of the manuscript. The corresponding author (FDL) attests that all listed authors meet authorship criteria and that no others meeting the criteria have been omitted.

